# Evaluating temperature and humidity gradients of COVID-19 infection rates in light of Non-Pharmaceutical Interventions

**DOI:** 10.1101/2020.07.20.20158071

**Authors:** Joshua Choma, Fabio Correa, Salah-Eddine Dahbi, Kentaro Hayasi, Benjamin Lieberman, Caroline Maslo, Bruce Mellado, Kgomotso Monnakgotla, Jacques Naudé, Xifeng Ruan, Finn Stevenson

## Abstract

We evaluate potential temperature and humidity impact on the infection rate of COVID-19 with a data up to June 10^*th*^ 2020, which comprises a large geographical footprint. It is critical to analyse data from different countries or regions at similar stages of the pandemic in order to avoid picking up false gradients. The degree of severity of NPIs is found to be a good gauge of the stage of the pandemic for individual countries. Data points are classified according to the stringency index of the NPIs in order to ensure that comparisons between countries are made on equal footing. We find that temperature and relative humidity gradients don’t significantly deviate from the zero-gradient hypothesis. Upper limits on the absolute value of the gradients are set. The procedure chosen here yields 6·10^−3 °^C^−1^ and 3.3·10^−3^ (%)^−1^ upper limits on the absolute values of the temperature and relative humidity gradients, respectively, with a 95% Confidence Level. These findings do not preclude existence of seasonal effects and are indicative that these are likely to be nuanced.

## 1. Introduction

In the last century the world has previously experienced three pandemics [1], the Asian, Spanish and Hong Kong influenza viruses. These pandemics had widespread consequences killing millions of people around the world [2, 3, 4]. There are many different factors that effect the transmission rate of a specific pandemic. These factors include, but are not limited to, climate change conditions, population density and medical quality [5]. The most recent pandemic is the novel COVID-19 Coronavirus caused by severe acute respiratory syndrome Coronavirus 2 (SARS-CoV-2) [6, 5]. The initial outbreak took place in December 2019 in Wuhan city, Hubei province, China [7]. This virus was declared pandemic by the World Health Organisation (WHO) on March 11, 2020 after it had spread into more than 114 countries with a total of 118,000 cases at the time [8]. As of June 18, 2020 the total number of cases identified by the WHO stands at around 8.2 million with fatalities reported at 445 thousands [9]. The spread of this virus is mainly based on human-to-human contact or droplets [10].

This paper aims to address the seasonality of the COVID-19 virus and whether it is driven by macroscopic temperature and/or humidity gradients. As the Southern Hemisphere has plunged into winter, concerns emerge regarding possible acceleration of the spread. Conversely, the Northern Hemisphere has transitioned to summer, where some countries appear confident that warmer weather will ease pressures from the pandemic.

A number of studies have been performed at different stages of the pandemic globally. Currently, the pandemic has reached a stage where many countries have completed their “first wave”. This wave cycle is defined by a slow initial growth, followed by an acceleration leading to an apex and then a gradual decline. However, the evolution of the pandemic does not occur in the same time frame for all countries. The length of the cycle and its initial parameters vary from country to country. If one analyses the infection rate in different countries at a fixed time it is possible to run into the error of comparing outcomes at different stages of the pandemic. For instance, at a given time a country can be at the early stages of the pandemic, which is characterized by low infection rates, while another can on be its way to reaching the apex with large infection rates. If both countries happen to display different temperatures or humidity levels, the analysis will yield an apparent gradient. Also, if one analyses infection rates over prolonged periods of time over which climatic conditions change significantly in a country, the analysis will also pick up an apparent gradient in the absence of genuine dependencies. The “first wave” in Europe commenced towards the end of the winter and was completed in the spring. At the time when the infection rates were large in some European countries in other continents, such as South America which happened to be warmer and more humid, displayed lower infection rates. From here One may be tempted to conclude on the presence of a gradient.

In order to reduce the probability of picking false gradients it is essential to compare infections rates for different countries at similar stages of the progression of the pandemic. Given the large geographical footprint of the pandemic attained so far it is possible to achieve a large enough differential in temperature and humidity in order to pin down potential macroscopic gradients. This is achieved here by classifying the data according to the level of severity of the non-pharmaceutical interventions (NPIs) applied in a country. We find that the severity of the total NPI policy is a good metric to gauge the progression of the pandemic. It must also be noted that countries with a number of positive cases below a certain threshold are not considered. This is motivated by the need to exclude countries that find themselves at the early stages of the pandemic, which is a difficult period to model. Several measures have been put in place by different countries in an attempt to limit the spread of this virus. Most countries around the world have adopted similar regulation related to combating the spread of COVID-19. Some of the most widespread strategies are: lockdown implementation, bans on mass gatherings, the closure of schools and work places and travel restrictions. A stringency index introduced by the Oxford COVID-19 Government Response Tracker team can be used to provide a picture of the stage at which any country enforced its strongest measures. The index is a number from 0 to 100 that reflects a weighted average of the inididual indicators indicators (related to each implemented NPI). A higher index score indicates a higher level of stringency.

In our work we are looking at countries with more than 5000 cases and thus far as of June 10, 2020 there are 65 countries that fit the criteria. Other human Corona viruses such as SARS-CoV and Middle East Respiratory Syndrome (MERS-CoV) survived for 2 - 9 days on different materials surfaces under high temperatures such as 30^°^ [11]. Their survival increased to 28 days under lower temperatures such as 4^°^. Additionally, SARS-CoV survives longer under lower temperatures and relative humidity, which could be the reason why countries with high temperature and relative humidity such as Malaysia, Indonesia and Thailand did not have major outbreaks of SARS-CoV in 2003 [12]. Several studies concluded that low temperature [13, 14] and low humidity favor the transmission of the virus [5, 15]. From this it is expected that the spread will reduce with a rise in both temperature and humidity. In contrast to these, there are studies which suggest that temperature does not play any significant role in the spread of COVID-19 [16, 17, 18]. Prata et al. [19] found a negative correlation between temperatures and number of daily cases of COVID-19 within the range 16.84^°^C to 25.84^°^C. However, the study shows that there is no evidence that there will be a decrease in the number of cases for temperatures above 25.84^°^C.

## 2. Non-Pharmaceutical Interventions and the Stringency Index

Non-pharmaceutical Interventions, or NPIs, are response measures put in place by a government to try and combat the spread of virus or infectious disease. There are a number of NPIs that most governments across the world have enforced. A global analysis of the impact of NPIs on the COVID-19 spread dynamics have shown that it is valuable to use a stringency index to classify these NPIs. The global analysis was done using the Oxford COVID-19 Government Response Team (OxCGRT) [20] stringency index as well as an adapted US stringency index that was designed by our team to be as similar as the OxCGRT index as possible for comparison purposes. The world average for the coefficient that linearises the level of transmission with respect to the OxCGRT stringency index is *α*_*s*_ = 0.01 ±0.0017 (95% C.I.) [21].

The OxCGRT stringency [22], p in our notation, can be understood as a number from 0-100 that is a weighted average of a number of relevant NPI indicators. The OxCGRT has created a database of NPI indicators and the corresponding dates of initiation. The stringency index is also included in this database. The OxCGRT stringency index can be useful in quantifying and characterising a country’s response to the COVID-19 pandemic. This makes it a useful comparative parameter. It can also be used to show the progression of a pandemic in a country and to determine an overall classification of a response as “tight” or “loose” control.

Figure 1 illustrates the impact of NPIs on the infection rate of the pandemic. The graph is obtained from data of 47 countries and states of the US before the release of lockdown restrictions. This graph bears witness of the strong correlation between the application of the NPIs and the control over the pandemic.

**Figure 1:**
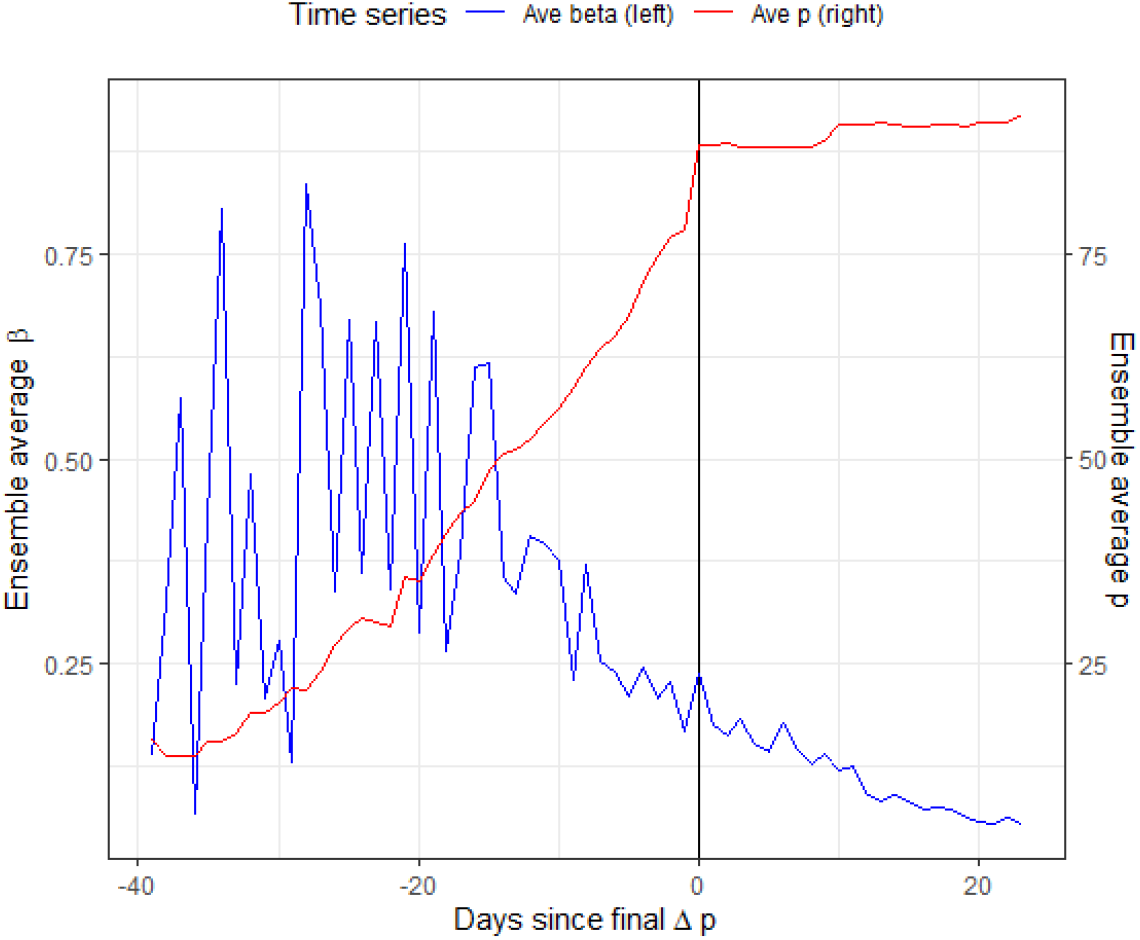
Comparison of ensemble average NPI stringency index, *p*, against the ensemble average of infection rates, β, as a function of days from the last major change in control value.

The OxCGRT original version that was used for this paper takes into account the following NPIs in the stringency index calculation:

1. S1 - School Closure
2. S2 - Workplace Closure
3. S3 - Cancel Public Events
4. S4 - Close Public Transport
5. S5 - Public Information Campaign
6. S6 - Domestic Travel Bans
7. S7 - International Travel Bans

The OxCGRT stringency index is given by:

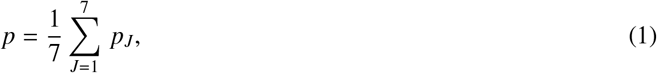

where *p*_*J*_ is defined by:

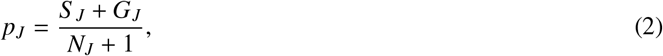

with *G*_*J*_ = 1 if the effect is general (and 0 otherwise), and *N*_*J*_ is the cardinality of the intervention measure [22, 20]. In the case where there is no requirement of general vs. targeted (S7), the +1 in the denominator and the *G*_*J*_ in the numerator are omitted from the equation to form:

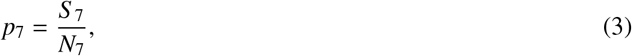

The OxCGRT database contains data for 133 countries however no data is included for US states. In order to be able to compare US states to countries, we mapped the known available levels of intervention in the US to match the OxCGRT system as accurately as possible. We used the Institute for Health Metrics and Evaluation (IHME) [23] dashboard to obtain six dates at which specific states imposed different NPIs. At the time of development the known NPIs for the US taken from the IHME dashboard were as follows:

Although some of the above US interventions were not directly comparable to the OxCGRT indicators, their individual impact on the stringency index was still valid and should be included in the calculation of the index. By including *U*1 and *U*3 with the appropriate weight into the calculation use for the OxCGRT index, an equivalent US index is created. The following equation was developed:

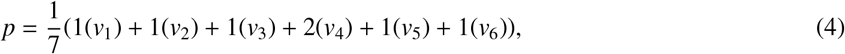

where *v*_*i*_ is a number out of 100 indicating the extent to which each of the interventions are imposed.

Due to lack of data on the *Travel Severely Limited* intervention on the IHME dashboard. It was required to source US travel restrictions information from other US news sources [24, 25, 26]. Using the same logic used by OxCGRT team the following equation for *v*_*i*_ was introduced:

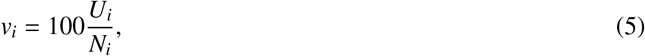

where *U*_*i*_ is ordinal and can vary from 0 to the cardinality of the specific intervention measure, *N*_*i*_. This is done to incorporate levels of implementation of a specific intervention into the stringency calculation. Based on the data the only intervention that requires levels of implementation is the *U*_4_ intervention. The ordinal levels, 0-3, were allocated for *U*_4_ in order to include the relevant levels of implementation that were found to be applicable to the *Travel Severely Limited* intervention.

## 3. Materials and Methods

### 3.1 The epidemiological parameters

Our study uses one of the simplest compartmental models in the form of SIRD modelling. The model consists of four compartments, *S, I, R* and *D* which represent the number of susceptible population, active cases, recoveries and number of deaths, respectively. The variables represent the number of people at the particular day in each compartment. The infection rate, *β*, the recovery rate, *γ*, and the mortality rate, *d* are defined. These parameters are given in the following formulae:

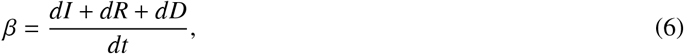

Where

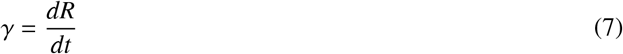

And

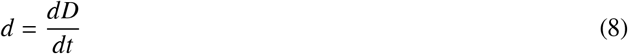

These parameters are computed on a daily basis. Figure 2 shows a snapshot of the infection rate globally for June 1^*st*^ 2020. A global analysis indicates that the recovery rate evolves slowly with time. In addition, the US stopped reporting recoveries early on in the pandemic. This data sample is particularly relevant to the analysis in that the differentials of temperature and humidity in the US are large. As a result, the temperature and humidity gradients are computed with respect to the infection rate, as opposed to the reproductive number.

**Figure 2:**
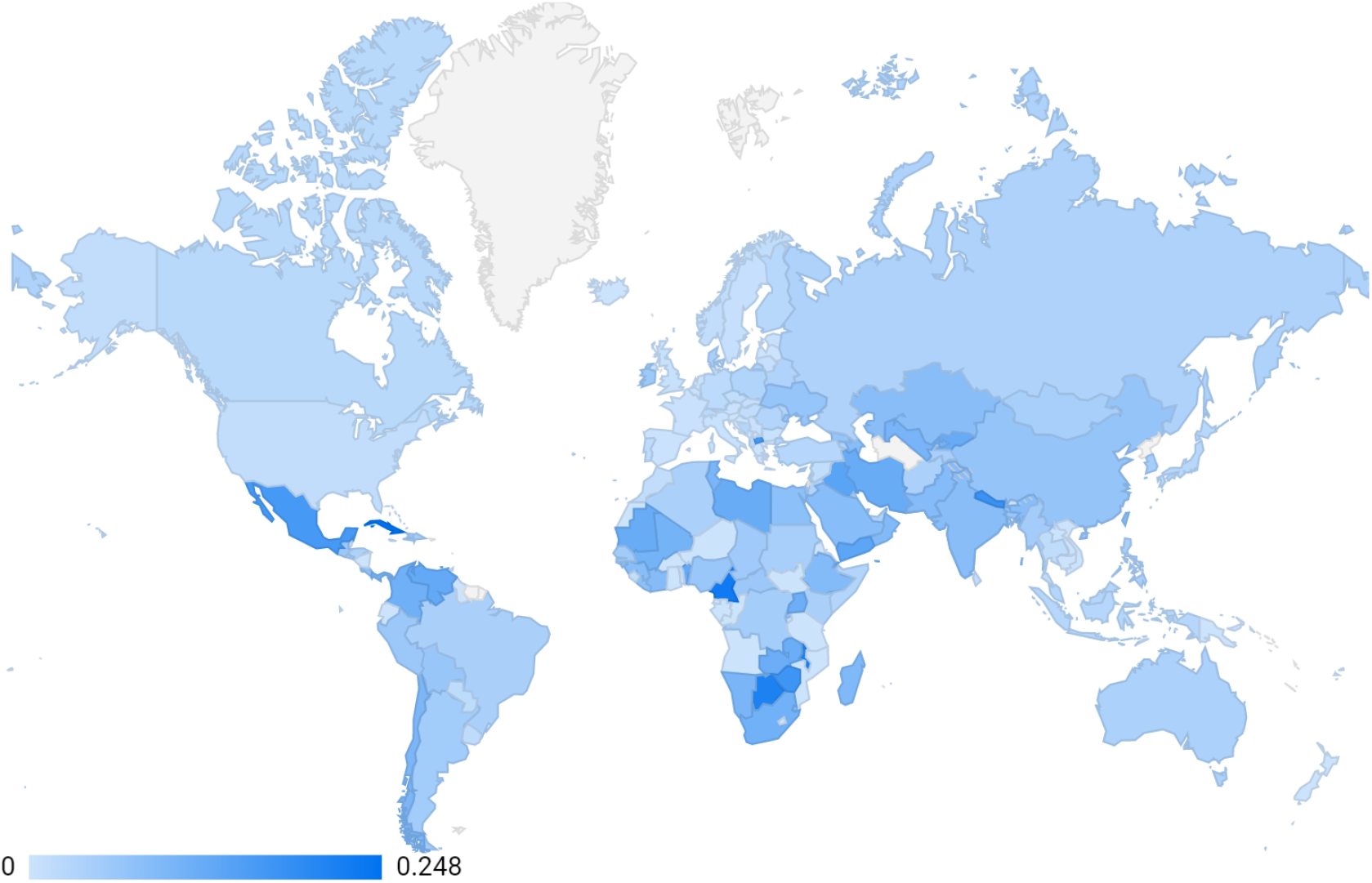
World map with daily *β* infection rate for the 1^*st*^ June 2020.

### 3.2 Disentangling climate modulations and the temporal evolution

As indicated in Section 1, the pandemic evolves in time undergoing different phases within a wave. A wave cycle can span months during which climate modulations may induce significant variations in temperature and humidity. Therefore, it is of the essence to make every effort to disentangle the intrinsic temporal evolution of a pandemic due to the specific characteristics of a country from climate modulations that may potentially impact it. This is a challenging exercise for a country in isolation. In the absence of a reliable overarching model, potential seasonality effects driven by regional temperature and humidity need to be extracted iteratively by means of a global analysis that later could be feed into a more localised scrutiny of the pandemic.

Should significant temperature and humidity gradients be impacting the dynamics of the pandemic at the macroscopic level, variations should be observed in a global and regional analysis of the data. In order to probe these potential gradients one has to compare epidemiological indexes in groups of countries that are at similar stages of the temporal evolution of the pandemic. This can be achieved by comparing infection rates as a function of the stringency index of the NPIs.

Most of the countries that have undergone the first gave went through at least three stages. Following the establishment of community transmission, a first stage in the pandemic is characterised by relatively large infection rates. At this point NPIs are not stringent or not sufficiently adhered to by the population. This state is referred to here as loose control. At this stage, the cumulative distribution of cases can be approximated by a single exponential. A second stage follows, where stringent NPIs are enforced. These measures are referred to here as tight control. The growth of cases decelerates, where the pandemic reaches an apex. The infection rates decrease more or less monotonically and, in many cases, the reproductive number drops below unity. Finally, a third stage follows, where Governments release lockdown measures more or less gradually with different levels of success.

As discussed in Section 1, an attempt is made here to avoid picking up false temperature and humidity gradients when comparing infection rates in countries or groups of countries that happen to be at different stages of the pandemic. A first iteration is performed here by evaluating temperature and humidity gradients in groups of countries that display “loose” or “tight” NPI control. This approach is effective as long the data displays sufficiently large differentials of temperature and humidity. In this light the data from the States of the US is an interest showcase, where progression of the pandemic displayed strong commonalities in the backdrop of large temperature and humidity differentials, e.g. the Midwest versus Florida and Arizona.

Given the differentials present in the data (see Section 4), this first iteration should be able to pick up gradients greater or comparable to 10^−2^. In order to refine the measurement of the gradient an approach more sophisticated than the discrete classification adopted here. For this purpose, parametric corrections would need to be implemented in the data to account for the stringency and the efficiency of the NPIs.

### 3.3 Data collection and processing

The study is done at the global level starting from January 24^*th*^ 2020 up to June 1^*st*^ 2020. The United States of America is considered separately because the stringency index is not available in the OxCGRT data base. The stringency index for the different states of the US is computed by the authors (see Section 2). This is done for the period January 25^*th*^ 2020 to April 14^*th*^, 2020.

The epidemiological data was collected from the Johns Hopkins University data repository which contains the global daily updates of the total cases, number of death and recovery cases [27]. In our study we focus on three cases. For the first case we consider all countries with more than 5000 total cases and US States with more than 2000 total cases. This is enforced in order to ensure that the epidemiological parameters be stable, as these are calculated on a daily basis.

For the second case, which is called loose control, we look at points where infection rate is not 0 and the stringency index is less than 50; lastly we have the tight control data sample, which focuses on the the period that starts two weeks after the maximum stringency index is achieved.

Table 2 shows the of countries with more than 5000 cases and US States with more than 2000. The number of data points before and after splitting into loose and tight NPI control are also given in Table 2.^1^

**Table 1:** Table Showing US Interventions Acquired from the IHME.

| Label | NPI |
| --- | --- |
| *U1*: | Stay at Home Order |
| *U2*: | Educational Facilities Closed |
| *U3*: | Non-essential Services Closed |
| *U4*: | Travel Severely Limited |
| *U5*: | Initial Workplace Closure |
| *U6*: | Banned Mass Gatherings |

**Table 2:** Number of data points for each of the three cases considered in this study.

|  | Countries with > 5000 Cases | No Split | Loose Control | Tight Control |
| --- | --- | --- | --- | --- |
| Asia | 18 | 1695 | 348 | 248 |
| Africa | 8 | 651 | 72 | 126 |
| Europe | 25 | 2124 | 411 | 336 |
| North America | 6 | 493 | 56 | 84 |
| South America | 7 | 572 | 51 | 98 |
| Oceania | 1 | 99 | 29 | 14 |
| Global | 65 | 5634 | 967 | 906 |
| USA | 22* | 1617 | 541 | 595 |

\*States with more than 2000 cases.

### 3.4 Temperature and Relative Humidity

The Integrated Surface Database (ISD) belonging to the National Oceanic and Atmospheric Administration (NOAA) consists of hourly and synoptic observations from over 35 000 stations worldwide [28]. These stations are identified by 11 character code made up of USAF (U.S. Air Force) and WBAN (Weather-Bureau-Army-Navy) fixed weather station identifiers in the database. The database contains different climate change parameters such as temperature, dew point, wind speed and sea level pressure, amongst others. Relevant data for this study, dew point, *T*_*d*_, and temperature, *T*, both measured in tenths of degrees Celsius (^°^C) was extracted using station identifiers and averaged per specific regions or countries [29]. The former indicates the quantity of moisture in the air whilst the latter indicate the air temperature. Other variables, such as saturated vapor pressure which measures the pressure applied by air mixed with water vapor [30], actual vapor pressure which measures the water vapor in a volume of air [31] and relative humidity are computed.

Vapor pressure (*E*) is calculated using temperature using the expression:

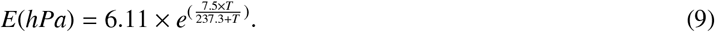

Saturated vapor pressure (*E*_*s*_) uses the dew point with:

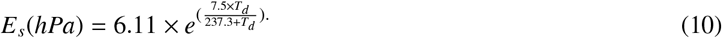

Here we use the relative humidity. This is given by the ratio of the two pressures and is measured in percentage:

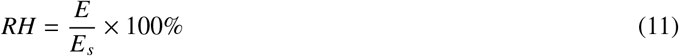

Figure 3 shows the global and USA maps with daily temperatures (left) and relative humidity (right) for June 1^*st*^ 2020.

**Figure 3:**
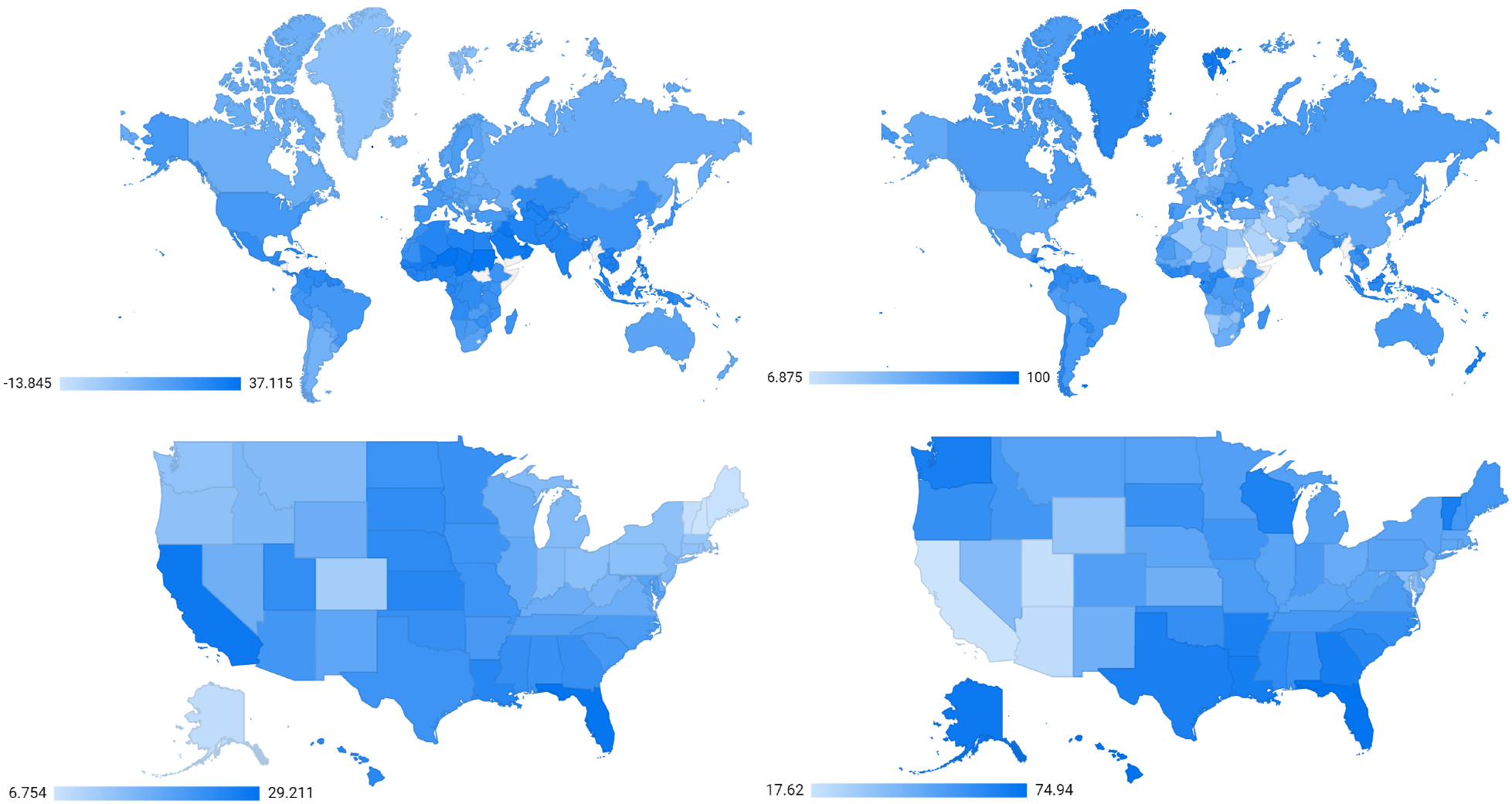
Maps of temperature (left) and relative humidity (right). Top plots depict global map, with USA being the bottom. Data correspond to June 1^*st*^ 2020.

## 4. Results and Discussion

The graphs in Figure 4 show the regression analysis of the transmission rate against temperature (top row) and relative humidity (bottom row) for the overall global data set discussed in Section 3. Results are shown for all three cases we have considered: the full data set, loose and tight controls, respectively. One appreciates the wide span of temperatures and relative humidity displayed by the data sample used here. The differential in relative humidity ranges from 90% to 80% for the full data set and the tight control sample, respectively. The corresponding differentials for temperature are 50 ^°^C and 44 ^°^C, respectively. Temperatures remain in the range between -15 ^°^C and 40 ^°^C. The differentials displayed in the data are sufficiently large to pick up gradients down to 10^−2^. For this purpose a regression analysis is performed with a first order polynomial of temperature and relative humidity. The available data does not seem to indicate that more complex functional forms are required. Statistical error propagation for the epidemiological parameters is performed.

**Figure 4:**
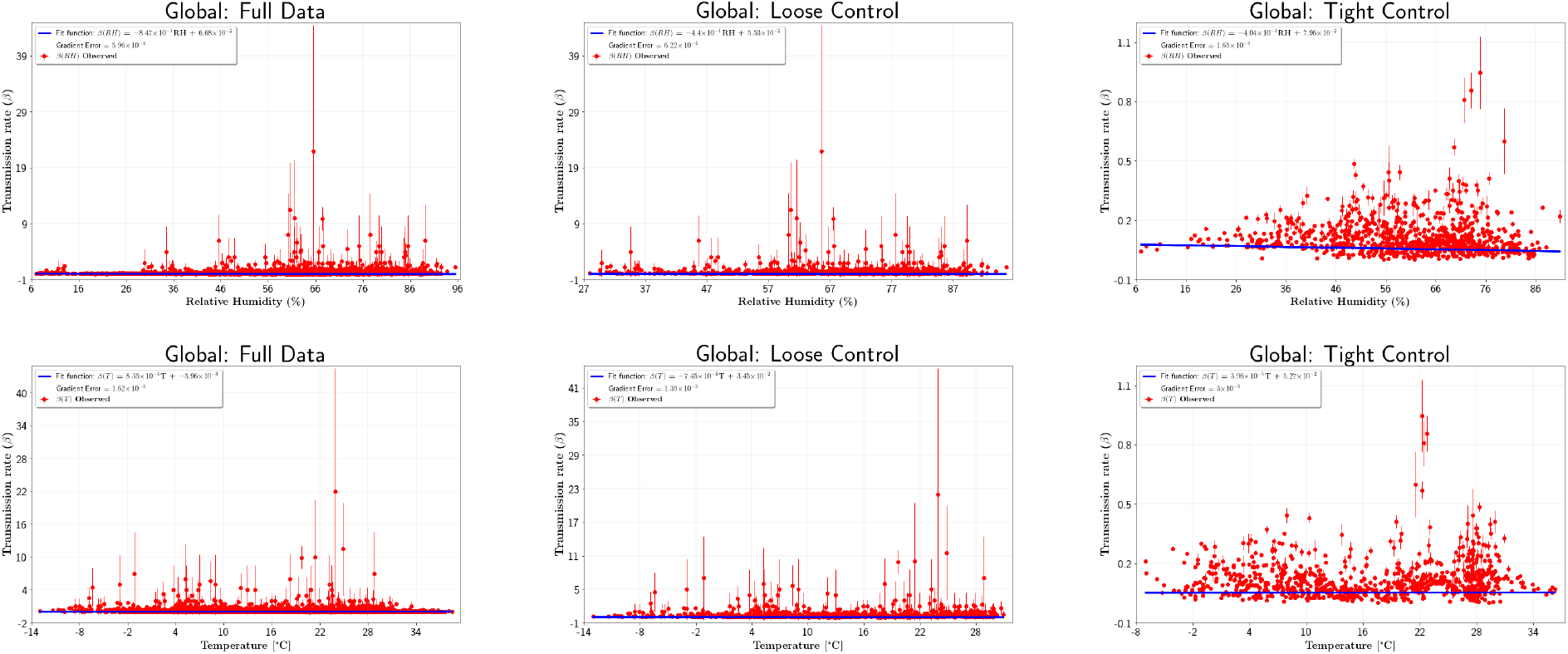
Regression analysis for relative humidity (top graphs) and temperature (bottom graphs) using global data, as detailed in Section 3. Results are shown for the full data set (left), loose control (centre) and tight control (right).

Tables 3 and 4 report the result of the regression analysis for the temperature and relative humidity analyses, respectively. Results are given in terms of central values and the 68% Confidence Intervals. The compatibility with the zero-gradient (absence of gradient) hypothesis is evaluated by means of a *p*-value analysis. Results from the global analysis indicate that the gradients are below 10^−3^ and that results are compatible with the zero-gradient hypothesis with a probability of better than 15%. These results indicate that the correlation between temperature and relative humidity with infection rates is weak.

**Table 3:** Temperature gradients with 68% Confidence Intervals and *p*-values for the zero-gradient hypothesis (see text).

|  | Full Data |  | Loose Control |  | Tight Control |  |
| --- | --- | --- | --- | --- | --- | --- |
| | Gradient ($^{\circ}\text{C}^{-1}$) | $p$-value | Gradient ($^{\circ}\text{C}^{-1}$) | $p$-value | Gradient ($^{\circ}\text{C}^{-1}$) | $p$-value |
| Asia | $2.17 \cdot 10^{-3} \pm 4.31 \cdot 10^{-4}$ | $< 0.001$ | $-4.17 \cdot 10^{-3} \pm 2.79 \cdot 10^{-3}$ | 0.136 | $1.70 \cdot 10^{-3} \pm 1.38 \cdot 10^{-3}$ | 0.218 |
| Africa | $-1.37 \cdot 10^{-3} \pm 5.61 \cdot 10^{-4}$ | 0.014 | $3.52 \cdot 10^{-3} \pm 2.90 \cdot 10^{-3}$ | 0.226 | $-1.45 \cdot 10^{-3} \pm 1.83 \cdot 10^{-3}$ | 0.427 |
| Europe | $-1.60 \cdot 10^{-3} \pm 1.91 \cdot 10^{-3}$ | 0.402 | $-9.96 \cdot 10^{-4} \pm 1.81 \cdot 10^{-3}$ | 0.581 | $-3.00 \cdot 10^{-3} \pm 4.52 \cdot 10^{-3}$ | 0.511 |
| North America | $3.80 \cdot 10^{-4} \pm 5.98 \cdot 10^{-4}$ | 0.525 | $4.33 \cdot 10^{-3} \pm 5.67 \cdot 10^{-4}$ | $< 0.001$ | $-1.32 \cdot 10^{-3} \pm 5.04 \cdot 10^{-4}$ | 0.008 |
| South America | $-3.51 \cdot 10^{-3} \pm 5.48 \cdot 10^{-3}$ | 0.005 | $-1.97 \cdot 10^{-2} \pm 2.78 \cdot 10^{-3}$ | $< 0.001$ | $-1.87 \cdot 10^{-3} \pm 4.06 \cdot 10^{-3}$ | 0.644 |
| Oceania | $1.49 \cdot 10^{-3} \pm 1.10 \cdot 10^{-3}$ | 0.177 | $-2.94 \cdot 10^{-2} \pm 7.08 \cdot 10^{-3}$ | $< 0.001$ | $1.55 \cdot 10^{-3} \pm 2.39 \cdot 10^{-3}$ | 0.516 |
| USA | $-5.75 \cdot 10^{-4} \pm 2.51 \cdot 10^{-3}$ | 0.819 | $2.32 \cdot 10^{-3} \pm 2.92 \cdot 10^{-3}$ | 0.427 | $-2.22 \cdot 10^{-3} \pm 3.70 \cdot 10^{-3}$ | 0.548 |
| Global | $8.32 \cdot 10^{-4} \pm 1.63 \cdot 10^{-3}$ | 0.607 | $-7.45 \cdot 10^{-4} \pm 1.39 \cdot 10^{-3}$ | 0.593 | $3.96 \cdot 10^{-5} \pm 3.00 \cdot 10^{-3}$ | 0.989 |

**Table 4:** Relative Humidity gradients with 68% Confidence Intervals and *p*-values for the zero-gradient hypothesis (see text).

|  | Full Data |  | Loose Control |  | Tight Control |  |
| --- | --- | --- | --- | --- | --- | --- |
| | Gradient ($\%^{-1}$) | $p$-value | Gradient ($\%^{-1}$) | $p$-value | Gradient ($\%^{-1}$) | $p$-value |
| Asia | $-2.71 \cdot 10^{-4} \pm 6.15 \cdot 10^{-4}$ | 0.660 | $-1.17 \cdot 10^{-3} \pm 6.49 \cdot 10^{-4}$ | 0.070 | $-5.66 \cdot 10^{-4} \pm 5.59 \cdot 10^{-4}$ | 0.312 |
| Africa | $-3.80 \cdot 10^{-4} \pm 1.57 \cdot 10^{-4}$ | 0.016 | $-3.17 \cdot 10^{-3} \pm 7.90 \cdot 10^{-4}$ | $< 0.001$ | $-5.56 \cdot 10^{-4} \pm 6.19 \cdot 10^{-4}$ | 0.370 |
| Europe | $-8.08 \cdot 10^{-4} \pm 5.72 \cdot 10^{-4}$ | 0.158 | $-2.33 \cdot 10^{-4} \pm 7.08 \cdot 10^{-4}$ | 0.742 | $-8.64 \cdot 10^{-4} \pm 1.96 \cdot 10^{-3}$ | 0.659 |
| North America | $-1.12 \cdot 10^{-3} \pm 7.35 \cdot 10^{-4}$ | 0.095 | $-4.96 \cdot 10^{-3} \pm 1.35 \cdot 10^{-3}$ | $< 0.001$ | $-2.34 \cdot 10^{-3} \pm 1.27 \cdot 10^{-3}$ | 0.067 |
| South America | $-3.35 \cdot 10^{-3} \pm 2.35 \cdot 10^{-3}$ | 0.155 | $-6.97 \cdot 10^{-3} \pm 2.11 \cdot 10^{-3}$ | $< 0.001$ | $3.19 \cdot 10^{-3} \pm 3.52 \cdot 10^{-3}$ | 0.365 |
| Oceania | $1.31 \cdot 10^{-3} \pm 5.16 \cdot 10^{-4}$ | 0.011 | $-3.57 \cdot 10^{-3} \pm 1.65 \cdot 10^{-3}$ | 0.030 | $1.87 \cdot 10^{-3} \pm 5.81 \cdot 10^{-4}$ | 0.001 |
| USA | $4.96 \cdot 10^{-4} \pm 3.71 \cdot 10^{-3}$ | 0.894 | $6.86 \cdot 10^{-3} \pm 4.46 \cdot 10^{-3}$ | 0.124 | $-2.24 \cdot 10^{-3} \pm 5.63 \cdot 10^{-3}$ | 0.691 |
| Global | $-8.47 \cdot 10^{-4} \pm 5.96 \cdot 10^{-4}$ | 0.155 | $-4.40 \cdot 10^{-4} \pm 6.22 \cdot 10^{-4}$ | 0.480 | $-4.04 \cdot 10^{-4} \pm 1.65 \cdot 10^{-3}$ | 0.807 |

The analysis is also performed for various continents, where the American continent is split into North, USA and South America. Results are also given in Tables 3 and 4 for temperature and relative humidity, respectively. In few instances the compatibility of the observed result with the zero-gradient hypothesis falls below 0.1%. However, this level of disagreement with the zero-gradient hypothesis is not observed consistently across the three data sets for any of the geographical regions considered here.

In the absence of conclusive evidence of temperature or humidity gradients in the data, upper limits on the absolute value of the gradients are set. These are set using the global data set with the tight control requirement. This choice yields more conservative limits compared to other data sets. This procedure yields 6·10^−3 °^C^−1^ and 3.3·10^−3^ (%)^−1^ upper limits on the absolute values of the temperature and relative humidity gradients, respectively, with a 95% Confidence Level.

The procedure followed here (see Section 3) is intended to capture the leading terms in the underlying dynamics driven by temperature or humidity. These terms appear small and seem compatible with the intrinsic uncertainties derived from reporting and epidemiological parameters. Given that the size of the terms are below the threshold of the analysis sensitivity, further implementation of more sophisticated functional forms of multi-dimensional approaches are not warranted here.

It is relevant to note that the weak correlation between temperature and humidity with the infection rate is observed in the full data set, even before the classification according to the severity of the NPIs is implemented. This seems to indicate that the size of the geographical footprint could play a significant role in removing biases. These biases could have been responsible for apparent gradients observed in studies performed with smaller data sets collected at earlier stages of the global pandemic.

## 5. Conclusions

Here we evaluate potential temperature and humidity impact on the infection rate of COVID-19. The data set used here comprises a large geographical footprint of data up to June 10^*th*^ 2020. This period covers the “first wave” of the pandemic in a large number of countries.

Our analysis indicates that it is critical to evaluate temperature and humidity gradients in different countries or regions at similar stages of the pandemic in order to avoid picking up false gradients. The degree of severity of NPI policy is found to be a good measure of the progression of the pandemic in a given country. As a result, data points are classified according to the stringency index of the NPIs in order to ensure that comparisons are made on equal footing.

We find that temperature and relative humidity gradients do not significantly deviate from the zero-gradient hypothesis. This implies that changes in temperature and relative humidity do not seem to have an effect on the value of the transmission rate or there is a small correlation between the transmission rate and temperature and relative humidity. Our results seem to be in agreement with other studies that has concluded that there is no evidence as to whether the spread of the COVID-19 is temperature dependent [16, 17, 18, 32].

In the absence of conclusive evidence of temperature or humidity gradients in the data, upper limits on the absolute value of the gradients are set. The procedure chosen here yields 6·10^−3 °^C^−1^ and 3.3·10^−3^ (%)^−1^ upper limits on the absolute values of the temperature and relative humidity gradients, respectively, with a 95% Confidence Level. The results obtained here speak to the absence of significant macroscopic temperature or humidity gradients. However, these results do not preclude the existence of seasonality effects in infection rates. It indicates that seasonal effects are likely to be nuanced and should not be reduced to temperature or humidity differentials.

## Data Availability

The COVID-19 stats data is available at the Johns Hopkins University data repository while the climate change data is from National Oceanic and Atmospheric Administration (NOAA).

https://www.ncei.noaa.gov/data/global-hourly/archive/csv/

## Footnotes

1 A data point is defined a vector of epidemiological parameters such that the estimated infection rate is not equal to zero. Data points are estimated on a daily basis.

